# Vestibular rehabilitation practice in neurorehabilitation: an Italian survey protocol

**DOI:** 10.1101/2024.04.15.24305831

**Authors:** Nicola Ferri, Jacopo Piermaria, Leonardo Manzari, Andrea Turolla, Giovanni Morone, Marco Tramontano

**Affiliations:** Department of Biomedical and Neuromotor Sciences (DIBINEM), Alma Mater Studiorum University of Bologna, Bologna, Italy; Division of Occupational Medicine, IRCCS University Hospital of Bologna S Orsola-Malpighi Polyclinic, Bologna, Italy; Santa Lucia Foundation IRCCS, Via Ardeatina 306, 00179 Rome, Italy; MSA ENT Academy Centre, Cassino, Italy; Department of Life, Health and Environmental Sciences, University of L’Aquila, 67100 L’Aquila, Italy

**Keywords:** Survey, Nervous System Diseases, Rehabilitation, video Head Impulse Test, Vestibular System, Exercise Therapy

## Abstract

**Introduction:** Neurological disorders are a prevalent cause of disability and death, which has a considerable impact on the world’s economic and healthcare systems, not least because of rehabilitation needs. Neurorehabilitation is effective and recommended for the best recovery of motor and cognitive functions and quality of life. This study evaluates clinicians’ perceptions of vestibular rehabilitation’s actual or potential role in neurorehabilitation.

**Methods:** This is the protocol for a cross-sectional study that will be conducted as an online survey approved by the University of Bologna Bioethics Committee. The survey lasts approximately 10 minutes and consists of 4 main sections (socio-demographic, professional, clinical, and future recommendation data), with a total of 27 closed questions and 2 open questions. 5 experts constructed the questionnaire, and then 6 respondents piloted it; these latter were representative of the subjects to whom it will be administered. We will report the Survey following the Checklist for Reporting Of Survey Studies (CROSS).

**Expected results:** This questionnaire will map the knowledge and use of vestibular rehabilitation in neurorehabilitation by healthcare professionals in Italy. It will also be useful to assess gaps, challenges, and research perspectives on this prevalent topic.

## Background

Neurological disorders are the first cause of disability and the second cause of death, with a worldwide estimate of one in three people being diagnosed at a certain point in their lifetime.^1^ Thus, it is a significant reason for rehabilitation needs, which does not end in the short term but could last for a long time in a patient’s life.^2^ This impacts the burden of neurological disorders on healthcare systems, representing an annual cost of 266 billion euros in Europe.^3^

Neurorehabilitation is a complex and multidisciplinary process effective in caring for people with nervous system injury, with the aim of improving the recovery of patients’ functionality and autonomy.^4^ There are several recommendations supporting neurorehabilitation to improve neuromotor and cognitive abilities in different neurological disorders like stroke,^5^ Parkinson’s disease,^6^ multiple sclerosis,^7^ concussion and mild traumatic brain injury.^8^

Vestibular rehabilitation is a treatment approach based mainly on exercises that target the postural and balance systems to recover from dysfunctions or injuries of the vestibular system.^9^ Recently, there has been a growing body of evidence supporting the effectiveness of vestibular rehabilitation in other neurological disorders as well. ^10–13^

A recent survey attempted to quantify vestibular knowledge among European therapists; however, Italy was not sufficiently represented, having included only one respondent.^14^ Thus, this study aims to map the knowledge, modalities, and perspectives of vestibular rehabilitation in persons with neurological disorders among Italian healthcare professionals, both at clinical and research levels.

## Methods

This online survey is a cross-sectional study in the Italian language aimed at healthcare professionals involved in vestibular rehabilitation. The Bioethics Committee at the University of Bologna has approved the study. The survey consists of four sections: sociodemographic information, professional background, clinical practice data, and future perspectives. A total of 27 closed and two open-ended questions were included.

Each section and each item were developed by a team of 5 experts, with an iterative feedback process to improve clarity, validity, fluency, and response time. Then, the draft was piloted by six independent respondents who are representative of our target population, and the final survey was obtained by implementing the feedback received. Our survey takes about 10 minutes to complete.

The demographic characteristics of respondents who piloted the surveys are reported in Table 1.

**Table 1.**
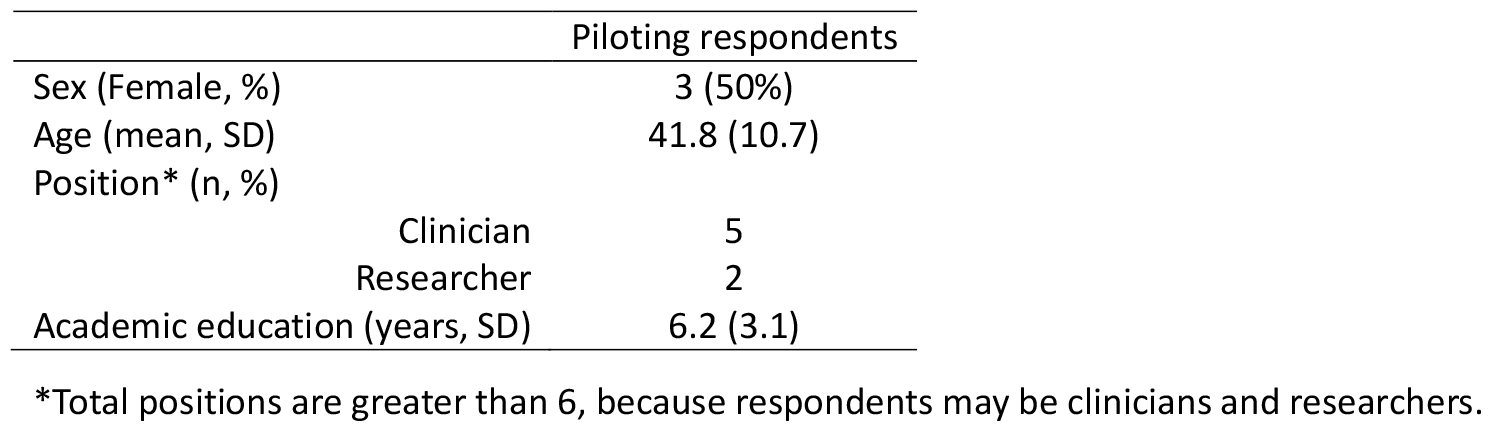
Piloting respondents (n= 6).

For our study, we defined vestibular rehabilitation as the set of exercises and strategies primarily targeting the vestibular system.

We followed methodological recommendations when conducting this research.^15,16^ The survey will be reported in accordance with the Checklist for Reporting Of Survey Studies (CROSS).^17^

The software will filter the respondents according to their IP address to avoid multiple answers from the same user. No personal identifying information will be collected; informed consent will be obtained online before the start of the survey.

The survey will be disseminated online via social channels (i.e., Facebook, X, LinkedIn) and through emails from national professional associations. We consider a minimum sample of 120 respondents to be appropriate. The survey will be open approximately from 22/04/2024 to 31/05/2024, and we’ll follow the Dillman approach^18^ to enhance survey compliance; in particular, we’ll send each week a reminder for three weeks. We will also repeatedly publish outlets via social media during the survey’s opening period.

Respondents will not be offered any incentive or compensation.

### Statistical analysis

Descriptive analyses will be presented to assess the respondents’ demographic characteristics and survey results. Tables and graphs will be reported to better understand the findings. Closed questions will be analyzed by frequencies for categorical variables and mean ± standard deviation for continuous variables. Open-ended questions will be coded by an independent researcher and reported in a thematic analysis. We will exclude completely blank surveys.

The survey and the output will be generated using Qualtrics software (Provo, UT, USA), and statistical analyses will be performed with STATA 18 software (StataCorp. 2023, College Station, TX, USA).

## Data Availability

All data produced in the present study are available upon reasonable request to the authors.

